# Impact of contact tracing on COVID-19 mortality: An impact evaluation using surveillance data from Colombia

**DOI:** 10.1101/2020.08.14.20158535

**Authors:** Andres I. Vecino-Ortiz, Juliana Villanueva Congote, Silvana Zapata Bedoya, Zulma M Cucunuba

## Abstract

**Background:** Contact tracing is a key part of the public health surveillance toolkit. However, it is labor intensive and costly to carry it out. Some countries have faced challenges implementing contact tracing, and no impact evaluations to our knowledge have assessed its impact on COVID-19 mortality. This study assesses the impact of contact tracing in a middle-income country and provides data to support the expansion of contact tracing strategies with the aim of improving infection control.

**Methods:** We obtained publicly available data on all confirmed COVID-19 cases in Colombia between March 2 and June 16, 2020. (N=54,931 cases over 135 days of observation). We proxied contact tracing performance as the proportion of cases identified through contact tracing out of all cases identified, as suggested by WHO guidelines. We calculated the daily proportion of cases identified through contact tracing across 37 geographical units (32 departments and five districts). Further, we used a sequential log-log fixed-effects model to estimate the 21-days, 28-days, 42-days and 56-days lagged impact of the proportion of cases identified through contact tracing on the daily number of COVID-19 deaths. Both the proportion of cases identified through contact tracing and the daily number of COVID-19 deaths are smoothed using 7-day moving averages. Models control for prevalence of active cases, second-degree polynomials, and mobility indices. Robustness checks to include supply-side variables were performed.

**Results:** We found that a 10 percent increase in the proportion of cases identified through contact tracing is related to COVID-19 mortality reductions between 0.8% and 3.4%. Our models explain between 47%-70% of the variance in mortality. Results are robust to changes of specification and inclusion of supply-side variables.

**Conclusion:** Contact tracing is instrumental to contain infectious diseases and its prioritization as a surveillance strategy will have a substantial impact on reducing deaths while minimizing the impact on the fragile economic systems of lower and middle-income countries. This study provides lessons for other LMIC.

## Introduction

Since the first case report on December 31, 2019 (1), the rapid spread of the novel coronavirus (SARS CoV-2) led to the worst pandemic in the century (2). In April 2020 over 2.4 million cases were detected and up to 165,000 deaths were reported worldwide. By mid-July, more than 14 million cases and 600,000 deaths were reported around the globe (3), being at that time the region of the Americas the most affected in the world with more than half of all cases. Specifically, in Latin America, poverty, a prevalent informal economy, inequality, and weak health and social service networks collided with the first wave of the pandemic (4). For this reason, the main challenge for all countries, but particularly for middle and lower-income ones (LMIC), is to contain the spread of the virus while minimizing the adverse consequences of lockdowns and other measures that threaten recent gains in terms of poverty reduction (5). This fact has led to calls for better instruments to strengthen epidemiological surveillance tools in order to break transmission chains and reduce the adverse consequences of both COVID-19 and non-pharmaceutical interventions (6). A key tool to break transmission chains is contact tracing (7), which has proved its effectiveness to control other deadly diseases such as the 2014 Ebola outbreak in West Africa (7,8).

In the specific case of COVID-19, many different strategies have emerged to improve contact tracing, ranging from Northern Ireland that has performed contact tracing through telephone calls (9) to South Korea and China which have used mobile phone apps (10). Previous models have suggested the high value that contact tracing can have on curbing the transmission (10-12).

However, contact tracing is extremely labor intensive and it is difficult to implement in most LMICs (5,13), particularly during peaks of the epidemic. Therefore it is likely to be left behind as a useful tool to curb outbreaks. For this reason, data on the effectiveness of contact tracing on mortality is key, so decision makers can make better informed decisions on whether including contact tracing within their priority setting processes.

Colombia’s political division is comprised by 37 geographical units, 32 departments and five districts (Santa Marta, Barranquilla, Buenaventura, Cartagena, and Bogota) located within those departments. In Colombia, contact tracing must be done jointly by both departmental/district and municipal governments. Departmental/district and local governments are in charge of identifying contacts, testing them and isolating them. The departmental heterogeneity on contact tracing performance represents an opportunity to assess the impact of contact tracing on mortality. The study aims at determining the extent to which differences in contact tracing explain changes in mortality in Colombia. This study will become a key resource for decision makers about the effectiveness of this tool and more broadly about how epidemiological surveillance can reduce the burden of COVID-19, particularly for LMIC.

## Materials and methods

### Data

In this study, we use publicly available and anonymized data on all confirmed novel Coronavirus cases obtained from the “Open Data” portal of the Colombian National Institute of Health. The de-identified data can be accessed here (14). This dataset was obtained on June 17^th^, 2020 and contains data on 54,931 cases identified between March 2^nd^ and June 16th, 2020 (135 days of observation period). This is our main sample. All novel Coronavirus cases are confirmed through Polymerase Chain Reaction in Real Time (PCR-RT). We included data for all 32 departments plus five districts for a total of 37 geographical units as described earlier. The main sample is used for our main analysis and one of the two robustness checks including the number of installed ICU beds per department (equations 1 and 2 below, respectively).

For the other robustness check, which includes the percentage of ICU occupation (equation 3 below), we took a different set of cases outside the study period of our main sample. All cases between June 17^th^ and July 15^th^ (study period of 28 days), for a total of 112,279 confirmed cases were obtained in July 17^th^, 2020 to run a robustness check including the percentage of ICU occupation by day and department to account for time-varying supply-side conditions. Percentage of ICU occupation was only available for that period of time and was obtained from the Ministry of Health dashboard (15). Cases for the robustness check double the original sample as a consequence of the exponential increase in COVID-19 cases in Colombia in mid-2020.

A useful performance metric to ascertain the completeness and quality of epidemiological surveillance is the proportion of cases that are identified through contact tracing out of all identified cases (16). In Colombia, cases identified through contact tracing are called “related cases”, which is a confirmed case detected through a test with a known epidemiological link (17). In this study, we use the 7-day moving average daily departmental proportion of related cases out of all cases detected as a proxy metric for contact tracing performance.

We used all COVID-19 confirmed deaths for the same period (N=1,853 for the main sample; N=3,738 for the robustness check sample), which is our outcome measure. To account for supply-side conditions, we also included data on total ICU beds per department. Unfortunately, we only have data on ICU beds installed as of June 14^th^, 2020 and data on ICU bed occupation between June 17^th^ and July 15^th^. Data on both ICU beds installed and percentage occupation of ICU beds is obtained from the Ministry of Health dashboard (15). We used data from the Google

Community Mobility Reports (18) for six modes of mobility: retail/recreation activities, trips to grocery/pharmacy stores, parks, transit stations and workplace sites. As the Google mobility report has data discriminated by department but not by district, we imputed the mobility data of the departments where the corresponding districts are located as follows: Santa Marta in Magdalena department; Barranquilla in Atlantico department; Buenaventura in Valle del Cauca department, and Cartagena in Bolivar department. The capital district of Bogota is included in Google as a separate department. The mobility index corresponds to the percentage change in mobility compared to the baseline, which is the median value for the corresponding day of the week, during the 5-week period between Jan 3-Feb 6, 2020. Finally, to capture concurrent time-varying epidemiological conditions that might affect surveillance (i.e. lack of capacity to surveil beyond a given number of cases) we calculated the number of daily active cases by department defined as the number of cases diagnosed minus those recovered or deceased. For the definition of active cases, we used the date of diagnosis. In the case of 622 cases from the main sample and 3,706 from the robustness check sample the date of diagnosis was not available. Therefore, we imputed the date of notification to the INS.

### Analysis

Our empirical strategy takes advantage of the heterogeneity of the proportion of cases that resulted from contact tracing, as a proxy of the effectiveness of contact tracing for each department over a 135-days observation period. We use a fixed-effects model, a widely accepted econometric technique to assess the effectiveness of public policy interventions that controls for both unobservable and observable time-invariant variables that might confound the relationship between contact tracing and mortality. With this identification approach, we prevent that baseline heterogeneity across departments (e.g. institutional strength of local health departments, funding, built capacity, etc.) bias our results.

We smoothed the trends of 1) daily departmental percentage of cases detected through contact tracing and 2) the daily number of COVID-19 deaths using a 7-day moving average. Next, we obtained logarithms for both metrics to be able to interpret the results as elasticities (percentage changes). Then, we assessed the relationship between the lagged (21, 28, 42 and 56 days) logarithm of the number of COVID-19 deaths and the logarithm of the percentage of the cases detected through contact tracing using a fixed-effects model and controlling for the logarithm of the number of active cases, for a second degree polynomial for contact tracing in order to account for any non-linear effects, and for the corresponding lag on the mobility index in six different dimensions of mobility.

As any effect that contact tracing has will only be observed at a population level between 2 and 6 weeks after (19), we obtained four different lags for the contact tracing variable at 21 days (model 1), 28 days (model 2), 42 days (model 3), and 56 days (model 4). We incrementally included lags to assess the effectiveness of contact tracing (sequential model).

In Equation 1 we present the full log-log fixed-effects model used in this study:

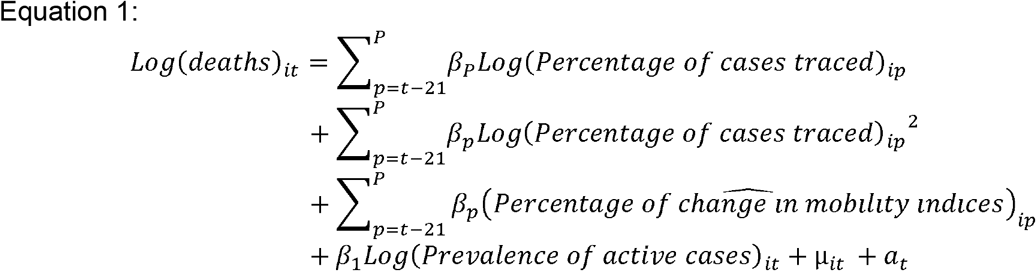

Where *i* is the department, *t* represents the day, β represents the coefficients, μ is the error term and a_i_ represents the department-specific intercept. p represents the four lagged periods observed: *t*-21; *t*-28; *t*-42; *t*-56.

### Robustness checks

Since supply-side conditions (e.g. ICU beds) are relevant for COVID-19 mortality. To capture supply-side effects, we used a random effects specification to introduce a time-invariant variable, the log of the number of ICU beds as of June 14^th^,2020. In Equation 2, we present the log-log random-effects model used in this study:

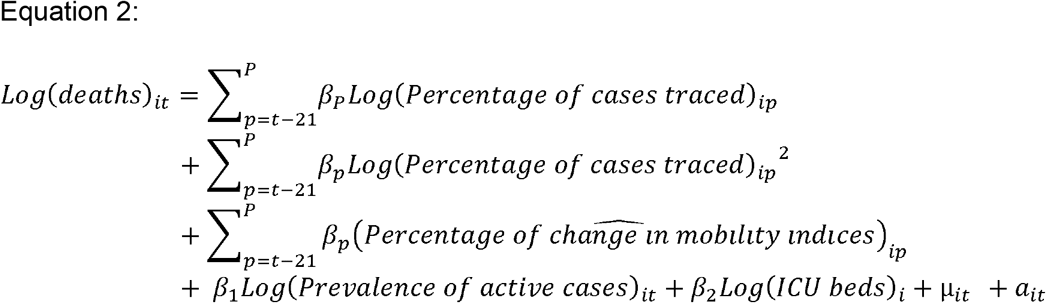

Where *i* is the department, *t* represents the day, β represents the coefficients, μ is the between-departments error term and a_it_ represents the within-department error term. p represents the four lagged periods observed: t-21; *t*-28; *t*-42; *t*-56.

Another way to capture the mortality effects of supply-side conditions is not only to ascertain the number of ICU installed beds as in Equation 2, but also the daily departmental percentage of ICU occupation. We obtained available data on percentage of ICU occupation by department between June 17^th^ and July 15^th^. We did not have data outside this period, but that fact allowed us to conduct this robustness check on a different set of cases. As the available period of study was 28 days, we ran model 1(21 days) and 2 (28 days) using a fixed effects model with the aim of capturing the time-varying effects of ICU on concurrent mortality. In Equation 3 we present the log-log fixed-effects model used in this study:

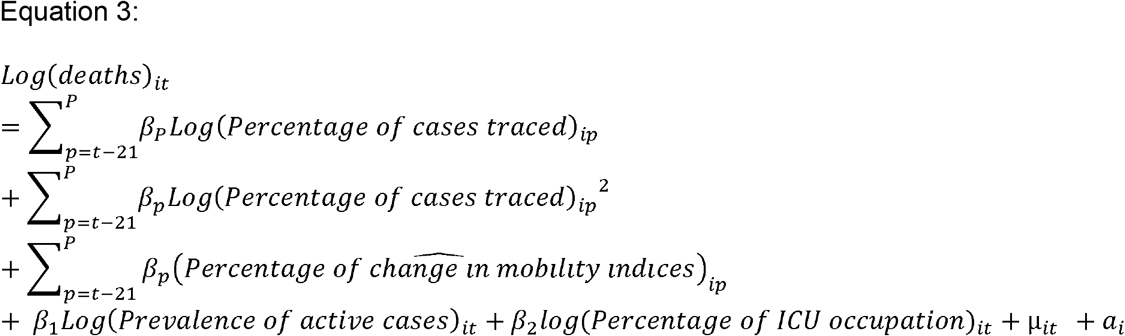

Where *i* is the department, *t* represents the day, β represents the coefficients, μ is the error term and a_i_ represents the department-specific intercept. p represents the two lagged periods observed: *t*-21; *t*-28.

## Results

From the 54,931 confirmed cases from March 2nd to June 16^th^, total daily cases by department ranged between 1 to 567 cases with a mean of 26.54 cases. Regarding the number of active cases the maximum was of 608 with a mean of 15.08, ICU beds per department ranged from 1 to 1206 with a mean of 276.6. The average of cases identified through contact tracing in our sample is 30.91% ranging between 0% to 100%. Total deaths by department by June 16 spanned between 1 to 24 with a mean of 2.65. Mobility indices averaged between −47% and −69% with overall mobility reduced across all departments. In table 1, we present these figures and also the figures for the sample for the robustness check.

**Table 1.** Descriptive statistics of the sample.

|  | Main sample |  |  | Sample for robustness check |  |  |
| --- | --- | --- | --- | --- | --- | --- |
|  | Mean | Minimum | Maximum | Mean | Minimum | Maximum |
| Total daily new cases by department | 26.54 | 1.00 | 567.00 | 113.01 | 12112.00 | 2112.00 |
| Number of active cases (incident cases) | 15.08 | -461.00 | 608.00 | 2064.56 | 338570.00 | 39238.00 |
| Number of ICUs beds per department | 276.50 | 1.00 | 1206.00 |  | 1.00 | 1206.00 |
| Percentage of ICU occupation by department |  |  |  | 56.12 | 0% | 10000% |
| Percentage of cases traced by department | 30.91% | 0% | 100% | 5.53% | 0% | 100% |
| Total deaths by department | 2.65 | 1.00 | 24.00 | 6.16 | 0.00 | 54.00 |
| Percentage change in mobility to retail/recreation activities | -67.68 | -93.00 | 5.00 |  | -89.00 | -24.00 |
| Percentage change in mobility to grocery/pharmacy stores | -47.19 | -91.00 | 32.00 |  | -82.00 | 10.00 |
| Percentage change in mobility in parks | -59.79 | -91.00 | 10.00 |  | -82.00 | -12.00 |
| Percentage change in mobility to transit stations | -69.25 | -100.00 | 17.00 |  | -100.00 | -6.00 |
| Percentage change in mobility to workplace sites | -46.96 | -89.00 | 25.00 |  | -79.00 | 7.00 |

Note: Main sample is comprised by 54,931 confirmed cases reported to the National Institute of Health. Surveillance data covers the period between March 2nd, 2020 and June 16th, 2020. Cases are defined as confirmed (PCR-RT) cases. Deaths include deaths of confirmed cases. Departments represent 32 departments plus 4 special districts (Santa Marta, Cartagena, Buenaventura and Barranquilla) and the capital district (Bogotá). Mobility data is obtained from Google and the values reflect percentage changes with regards to the baseline. the data corresponds on the mobility index in six different modes of mobility: retail/recreation activities, trips to grocery/pharmacy stores, parks, transit stations and workplace sites. The mobility index corresponds to the percentage change with regards to the baseline, which is the median value for the corresponding day of the week, during the 5-week period Jan 3-Feb 6, 2020.
Sample for robustness checks is comprised by 112,279 confirmed cases reported to the National Institute of Health between June 17, 2020 and July 15, 2020 for a 28-day observation period. Departments represent 26 departments plus 4 special districts (Santa Marta, Cartagena, Buenaventura and Barranquilla) and the capital district (Bogotá). We excluded the departments of Amazonas, Choco, Guaviare, Guainia, Vaupes, and Vichada because they did not have consistent data on ICU occupation in the 28-day sub study period.

As it can be seen in Table 2, performance metrics for contact tracing in the previous 21 to 56 days significantly predict mortality reductions in Colombia. An increase in 10% in the proportion of cases identified through contact tracing is related to mortality reductions in the next 21 to 56 days of between 2.1 and 2.3%, depending on the model. Second-order polynomials reveal a slight but statistically significant convex behavior. As these results are from cases obtained before the surveillance system is overwhelmed, it is possible that as cases increases, the second derivative turns concave and marginally decreasing returns might take place.

**Table 2.** Fixed effects design at the department level on the effect of contact tracing on COVID-19 mortality.

| Log of the number of COVID-19 deaths | Model 1<br>(lag for 21 days) |  |  | Model 2<br>(lags for 21 and 28 days) |  |  | Model 3<br>(lags for 21, 28, and 42 days) |  |  | Model 4<br>(lags for 21, 28, 42, and 56 days) |  |  |
| --- | --- | --- | --- | --- | --- | --- | --- | --- | --- | --- | --- | --- |
|  | Effect | SE | *p* | Effect | SE | *p* | Effect | SE | *p* | Effect | SE | *p* |
| Logarithm of the percentage of cases traced 21 days before | -0.212 | 0.036 | <0.001 | -0.210 | 0.038 | <0.001 | -0.212 | 0.039 | <0.001 | -0.190 | 0.040 | <0.001 |
| Logarithm of the percentage of cases traced 28 days before |  |  |  | -0.011 | 0.033 | 0.738 | -0.001 | 0.034 | 0.981 | -0.010 | 0.035 | 0.782 |
| Logarithm of the percentage of cases traced 42 days before |  |  |  |  |  |  | -0.003 | 0.029 | 0.929 | 0.005 | 0.030 | 0.868 |
| Logarithm of the percentage of cases traced 56 days before |  |  |  |  |  |  |  |  |  | -0.032 | 0.025 | 0.211 |
| Squared of the logarithm of the percentage of cases traced 21 days before | 0.009 | 0.002 | 0.844 | 0.009 | 0.002 | <0.001 | 0.009 | 0.002 | <0.001 | 0.008 | 0.002 | <0.001 |
| Squared of the logarithm of the percentage of cases traced 28 days before |  |  |  | 0.001 | 0.001 | 0.409 | 0.000 | 0.001 | 0.789 | 0.001 | 0.001 | 0.244 |
| Squared of the logarithm of the percentage of cases traced 42 days before |  |  |  |  |  |  | 0.001 | 0.001 | 0.174 | 0.001 | 0.001 | 0.014 |
| Squared of the logarithm of the percentage of cases traced 56 days before |  |  |  |  |  |  |  |  |  | 0.001 | 0.001 | 0.056 |
| R-square (within) |  | 0.503 |  |  | 0.523 |  |  | 0.512 |  |  | 0.507 |  |
| R-square (between) |  | 0.519 |  |  | 0.399 |  |  | 0.428 |  |  | 0.438 |  |
| Observations (department/day units) |  | 549 |  |  | 542 |  |  | 521 |  |  | 497 |  |

Note: N=54,931 confirmed cases reported to the National Institute of Health. The outcome is the logarithm of the daily number of COVID-19 deaths (7-day moving average) mortality rate between March 2nd, 2020 and June 16th, 2020. Cases are defined as confirmed (PCR-RT) cases. Deaths include deaths of confirmed cases. Departments represent 32 departments plus 4 special districts (Santa Marta, Cartagena, Buenaventura and Barranquilla) and the capital district (Bogotá). All models represented are a fixed effects model with intercepts for department. All models control for the logarithm of the 7-day moving average of new active cases in each department, for second-degree polynomials of the percentage of cases detected through contact tracing, and for the corresponding lag on the mobility index in six different modes of mobility: retail/recreation activities, trips to grocery/pharmacy stores, parks, transit stations and workplace sites. The mobility index corresponds to the percentage change with regards to the baseline, which is the median value for the corresponding day of the week, during the 5-week period Jan 3-Feb 6, 2020. The coefficients presented represent the logarithm of the percentage of cases traced (7-day moving average). Stratification is done for 21, 28, 42 and 56 days in advance of the observed date.

Seemingly, in table 3 we display the results from the random-effects design where the number of ICU beds per department were included with no major changes with respect to the fixed-effects model and a range of reduction in mortality between 2.1% (model 1) and 3.4% (model 3) after a 10% increase on the proportion of cases detected through contact tracing.

**Table 3.** Random effects design at the department level on the effect of contact tracing on COVID-19 mortality.

| Log of the number of COVID-19 deaths | Model 1<br>(lag for 21 days) |  |  | Model 2<br>(lags for 21 and 28 days) |  |  | Model 3<br>(lags for 21, 28, and 42 days) |  |  | Model 4<br>(lags for 21, 28, 42, and 56 days) |  |  |
| --- | --- | --- | --- | --- | --- | --- | --- | --- | --- | --- | --- | --- |
|  | Effect | SE | *p* | Effect | SE | *p* | Effect | SE | *p* | Effect | SE | *p* |
| Logarithm of the percentage of cases traced 21 days before | -0.217 | 0.035 | <0.001 | -0.214 | 0.037 | 0.074 | -0.229 | 0.041 | <0.001 | -0.203 | 0.042 | <0.001 |
| Logarithm of the percentage of cases traced 28 days before |  |  |  | -0.014 | 0.032 | 0.656 | 0.030 | 0.035 | 0.384 | 0.012 | 0.036 | 0.737 |
| Logarithm of the percentage of cases traced 42 days before |  |  |  |  |  |  | -0.067 | 0.030 | 0.024 | -0.062 | 0.030 | 0.040 |
| Logarithm of the percentage of cases traced 56 days before |  |  |  |  |  |  |  |  |  | -0.080 | 0.026 | 0.002 |
| Squared of the logarithm of the percentage of cases traced 21 days before | 0.008 | 0.002 | 0.478 | 0.009 | 0.002 | <0.001 | 0.010 | 0.002 | <0.001 | 0.009 | 0.002 | <0.001 |
| Squared of the logarithm of the percentage of cases traced 28 days before |  |  |  | 0.001 | 0.001 | -0.001 | 0.001 | 0.002 | 0.327 | -0.001 | 0.001 | 0.701 |
| Squared of the logarithm of the percentage of cases traced 42 days before |  |  |  |  |  |  | 0.001 | 0.001 | 0.205 | 0.002 | 0.001 | 0.058 |
| Squared of the logarithm of the percentage of cases traced 56 days before |  |  |  |  |  |  |  |  |  | 0.001 | 0.001 | 0.171 |
| R-square (within) |  | 0.503 |  |  | 0.521 |  |  | 0.468 |  |  | 0.449 |  |
| R-square (between) |  | 0.519 |  |  | 0.543 |  |  | 0.677 |  |  | 0.747 |  |
| Observations (department/day units) |  | 549 |  |  | 542 |  |  | 521 |  |  | 497 |  |

Note: N=54,931 confirmed cases reported to the National Institute of Health. The outcome is the logarithm of the daily number of COVID-19 deaths (7-day moving average) mortality rate between March 2nd, 2020 and June 16th, 2020. Cases are defined as confirmed (PCR-RT) cases. Deaths include deaths of confirmed cases. Departments represent 32 departments plus 4 special districts (Santa Marta, Cartagena, Buenaventura and Barranquilla) and the capital district (Bogotá). All models represented are a fixed effects model with intercepts for department. All models control for the logarithm of the 7-day moving average of new active cases in each department, the logarithm of the number of ICU beds in that department as of June 15th to account for supply-side factors, for second-degree polynomials of the percentage of cases detected through contact tracing, and for the corresponding lag on the mobility index in six different modes of mobility: retail/recreation activities, trips to grocery/pharmacy stores, parks, transit stations and workplace sites. The mobility index corresponds to the percentage change with regards to the baseline, which is the median value for the corresponding day of the week, during the 5-week period Jan 3-Feb 6, 2020. The coefficients presented represent the logarithm of the percentage of cases traced (7-day moving average). Stratification is done for 21, 28, 42 and 56 days in advance of the observed date.

In table 4, we present the results of the robustness checks including percentage of ICU occupation with a different set of cases, where an increase in 10% in the proportion of cases identified through contact tracing is related to mortality reductions in the next 21 to 28 days of between 0.8% and 2.2%, depending on the model.

**Table 4.** Fixed effects design at the department level on the effect of contact tracing on COVID-19 mortality.

| Log of the number of COVID-19 deaths | Model 1<br>(lag for 21 days) |  |  | Model 2<br>(lags for 21 and 28 days) |  |  |
| --- | --- | --- | --- | --- | --- | --- |
|  | Effect | SE | *p* | Effect | SE | *p* |
| Logarithm of the percentage of cases traced 21 days before | -0.078 | 0.034 | 0.024 | -0.082 | 0.034 | 0.017 |
| Logarithm of the percentage of cases traced 28 days before |  |  |  | -0.139 | 0.039 | <0.001 |
| Squared of the logarithm of the percentage of cases traced 21 days before | 0.000 | 0.001 | 0.991 | 0.000 | 0.002 | 0.012 |
| Squared of the logarithm of the percentage of cases traced 28 days before |  |  |  | 0.004 | 0.019 | 0.170 |
| R-square (within) |  | 0.307 |  |  | 0.351 |  |
| R-square (between) |  | 0.665 |  |  | 0.680 |  |
| Observations (department/day units) |  | 383 |  |  | 383 |  |

## Conclusion

This study is the first to our knowledge identifying substantial mortality reductions associated to contact tracing for COVID-19 and provides guidance on its effectiveness not only for Colombia but also potentially for other LMICs. An increase in 10% of cases detected through contact tracing in the 3 to 8 weeks before might be related to reductions in mortality ranging between 0.8% and 3.4%

Contact tracing is a known infection containment strategy recommended by the World Health Organization (16). Several modeling studies have suggested contact tracing would allow easing lockdown restrictions, particularly in LMICs with high levels of informality and weak health and social services networks (10). Rapid identification, and isolation of COVID-19 contacts increases detection of secondary cases and avoids the spread to tertiary cases, which might have an impact on mortality and ICU occupation.

Contact tracing capabilities often rely on strong public health infrastructure, well-trained human resources and adequate funding, which in many LMIC’s might be lacking, especially in rural areas.

One of the many challenges of contact tracing is keeping track of the contacts as the epidemic grows. The ability for tracing enough contacts in a timely fashion (11) is substantially better when contact rates are lower. Previous modelling work suggests that there is a strong synergic effect between contact rates and ability of contact tracing, primarily mediated by the traceability of those (20).

Traditional strategies for contact tracing have always relied on trained staff, which many times include volunteers, students and retirees, as well as health professionals, NGO workers, and community members (6). They carry out tracking activities that include locating and identifying infectious case contacts, and with coordinating support, allow for the monitoring and management of information to guide decision-making. Another strategy is community-based contact tracing which allows for improved community engagement and ownership of this epidemiological surveillance measures (21).

Finally, there are technology-based contact tracing strategies that take advantage of mobile phone apps to identify proximity of cases. From a public health perspective, these technologies are highly useful, but they often present challenges including privacy concerns and limitations on mobile network coverage.

This paper presents some limitations. 1) We are using a performance parameter to proxy contact tracing, which might be endogenously related to the performance in non-pharmaceutical interventions. However, we control for mobility indices with the aim of reducing potential endogeneity due to concurrent lockdowns. 2) For our metric of contact tracing to capture mortality effects, needs to be accompanied by isolation of the contacts. As we cannot measure isolation performance from this data, the assumption is that there is homogeneous isolation compliance across departments. This is important because contact tracing is not effective without effective isolation and our indicator might represent a broader indicator for epidemiological surveillance performance rather than only contact tracing. 3) Officially confirmed COVID deaths might be underestimated as some suspected cases might not be classified as COVID-19 deaths. A recent report of the Ministry of Health flagged this issue (22), but no specific figures on COVID-19 suspected mortality have been provided at this time. However, the ministry estimates an overall excess mortality of only 3.8% for the first semester of 2020. 4) We are providing data only for Colombia because of the quality of the datasets that can be obtained. However, we expect that the lessons of this paper can guide decision-makers in other LMICs about the impact that contact tracing can have on reducing COVID-19 mortality while also allowing less restrictive or more localized lockdowns to minimize the adverse economic and societal impacts these have.

Our study suggests contact tracing is effective in reducing mortality. Nonetheless, no epidemiological surveillance strategy should be isolated from other measures. Whereas we are measuring a performance metric for contact tracing, other tools such as increased testing, isolation, and economic and social support for contacts must be part of an integrated approach.

## Data Availability

The anonymized surveillance data used in this study is publicly available at https://www.datos.gov.co/Salud-y-Protecci-n-Social/Casos-positivos-de-COVID-19-en-Colombia/gt2j-8ykr/data.

## Acknowledgements

We thank the Colombian National Institute of Health for making this data available for research and input.

## Contributions to the literature

- Contact tracing is a key part of the public health surveillance toolkit. However, there are no previous statistical inference studies assessing the effectiveness of contact tracing in reducing mortality.
- Contact tracing is effective in reducing mortality even when controlling for mobility indices (as a proxy for lockdown policies) and supply-side factors.
- Contact tracing changes have a relevant effect on mortality for COVID-19 as soon as three weeks after implemented.

## References

1. Koo JR, Cook AR, Park M, Sun Y, Sun H, Lim JT, et al. Interventions to mitigate early spread of SARS-CoV-2 in Singapore: a modelling study. Lancet Infect Dis. 2020;20(6):678–88.

2. Tang Y, Wang S. Mathematic modeling of COVID-19 in the United States. Emerg Microbes Infect. 2020 Dec;9(1):827-9.

3. Dong E, Du H, Gardner L. An interactive web-based dashboard to track COVID-19 in real time. Lancet Infect Dis. 2020;20(5):533–4.

4. Burki T. COVID-19 in Latin America. The Lancet Infectious Diseases. 2020 May 1;20(5):547–8.

5. Walker PGT, Whittaker C, Watson OJ, Baguelin M, Winskill P, Hamlet A, et al. The impact of COVID-19 and strategies for mitigation and suppression in low- and middle-income countries. Science [Internet]. 2020 Jun 12 [cited 2020 Jun 27]; Available from: https://science.sciencemag.org/content/early/2020/06/11/science.abc0035

6. World Health Organization. Surveillance strategies for COVID-19 human infection [Internet]. 2020 [cited 2020 Jun 27]. Available from: https://www.who.int/publications-detail-redirect/surveillance-strategies-for-covid-19-human-infection

7. World Health Organization. Contact Tracing During an Outbreak of Ebola Virus Disease [Internet]. 2014. Available from: https://www.who.int/csr/resources/publications/ebola/contact-tracing-during-outbreak-of-ebola.pdf?ua%20=%201

8. World Health Organization. WHO | Implementation and management of contact tracing for Ebola virus disease [Internet]. World Health Organization; 2015 [cited 2020 Jun 24]. Available from: http://www.who.int/csr/resources/publications/ebola/contact-tracing/en/

9. Lee VJ, Chiew CJ, Khong WX. Interrupting transmission of COVID-19: lessons from containment efforts in Singapore. J Travel Med [Internet]. 2020 May 18 [cited 2020 Jun 27];27(3). Available from: https://academic.oup.com/jtm/article/27/3/taaa039/5804843

10. Ferretti L, Wymant C, Kendall M, Zhao L, Nurtay A, Abeler-Dörner L, et al. Quantifying SARS-CoV-2 transmission suggests epidemic control with digital contact tracing. Science [Internet]. 2020 May 8 [cited 2020 Jun 24];368(6491). Available from: https://science.sciencemag.org/content/368/6491/eabb6936

11. Kretzschmar ME, Rozhnova G, Bootsma MCJ, Boven M van, Wijgert JHHM van de, Bonten MJM. Impact of delays on effectiveness of contact tracing strategies for COVID-19: a modelling study. The Lancet Public Health [Internet]. 2020 Jul 16 [cited 2020 Jul 20];0(0). Available from: https://www.thelancet.com/journals/lanpub/article/PIIS2468-2667(20)30157-2/abstract

12. Keeling MJ, Hollingsworth TD, Read JM. Efficacy of contact tracing for the containment of the 2019 novel coronavirus (COVID-19). J Epidemiol Community Health [Internet]. 2020 Jun [cited 2020 Aug 13]; Available from: https://www.ncbi.nlm.nih.gov/pmc/articles/PMC7307459/

13. Heymann DL, Shindo N. COVID-19: what is next for public health? The Lancet. 2020 Feb 22;395(10224):542–5.

14. Open data platform of the Colombian Government. Casos positivos de COVID-19 en Colombia | Datos Abiertos Colombia [Internet]. la plataforma de datos abiertos del gobierno colombiano. 2020 [cited 2020 Jun 24]. Available from: https://www.datos.gov.co/Salud-y-Protecci-n-Social/Casos-positivos-de-COVID-19-en-Colombia/gt2j-8ykr/data

15. Ministerio de Salud y Protección Social. ArcGIS Dashboards [Internet]. 2020 [cited 2020 Jun 27]. Available from: https://minsalud.maps.arcgis.com/sharing/rest/oauth2/authorize?client_id=opsdashboard&display=default&response_type=token&expiration=20160&redirect_uri=https%3A%2F%2Fminsalud.maps.arcgis.com%2Fapps%2Fopsdashboard%2FpostSignIn.html&locale=en-us&state=%7B%22redirect%22%3A%22https%3A%2F%2Fminsalud.maps.arcgis.com%2Fapps%2Fopsdashboard%2Findex.html%22%2C%22portalUrl%22%3A%22https%3A%2F%2Fminsalud.maps.arcgis.com%2Fsharing%2Frest%2F%22%7D

16. World Health Organization. Contact tracing in the context of COVID-19 [Internet]. 2020 [cited 2020 Jun 24]. Available from: https://www.who.int/publications-detail-redirect/contact-tracing-in-the-context-of-covid-19

17. Instituto Nacional de Salud. Instructivo para la vigilancia en salud pública intensificada de infección respiratoria aguda asociada al nuevo coronavirus 2019 [Internet]. 2020. Available from: https://www.ins.gov.co/Noticias/Coronavirus/Anexo_%20Instructivo%20Vigilancia%20COVID%20v11%2012052020.pdf

18. Google. COVID-19 Community Mobility Report [Internet]. COVID-19 Community Mobility Report. 2020 [cited 2020 Jun 30]. Available from: https://www.google.com/covid19/mobility?hl=en

19. Gaythorpe K, Imai N, Cuomo-Dannenburg G, Baguelin M, Bhatia S, Boonyasiri A, et al. Report 8: Symptom progression of COVID-19 [Internet]. 11. 2020 Mar [cited 2020 Jun 24]. Available from: http://spiral.imperial.ac.uk/handle/10044/1/77344

20. Kucharski AJ, Klepac P, Conlan AJK, Kissler SM, Tang ML, Fry H, et al. Effectiveness of isolation, testing, contact tracing, and physical distancing on reducing transmission of SARS-CoV-2 in different settings: a mathematical modelling study. The Lancet Infectious Diseases [Internet]. 2020 Jun 16 [cited 2020 Jun 30];0(0). Available from: https://www.thelancet.com/journals/laninf/article/PIIS1473-3099(20)30457-6/abstract

21. Wong V, Cooney D, Bar-Yam Y. Beyond Contact Tracing: Community-Based Early Detection for Ebola Response. PLoS Curr [Internet]. 2016 May 19 [cited 2020 Jun 27];8. Available from: https://www.ncbi.nlm.nih.gov/pmc/articles/PMC4946441/

22. Ministerio de Salud y Protección Social. EXCESO DE MORTALIDAD EN COLOMBIA 2020 [Internet]. 2020. Available from: https://www.minsalud.gov.co/sites/rid/Lists/BibliotecaDigital/RIDE/VS/ED/VSP/estimacion-exceso-mortalidad-Colombia-2020.pdf

